# Polygenic Background Contributes to GCK-MODY Clinical Presentation and Glycaemic Variability

**DOI:** 10.1101/2025.08.04.25332935

**Authors:** Jacques Murray Leech, Ankit M Arni, V. Kartik Chundru, Luke N Sharp, Kevin Colclough, Andrew T Hattersley, Michael N Weedon, Kashyap A Patel

**Affiliations:** Department of Clinical and Biomedical Sciences, Faculty of Health and Life Sciences, University of Exeter, Exeter, UK; Exeter Genomics Laboratory, Royal Devon University Healthcare NHS Foundation Trust, Exeter, UK

**Keywords:** Diabetes, Genetics, Genetic Risk, Glucokinase, HbA1c, Maturity-onset diabetes of the young, MODY, Monogenic Diabetes, Polygenic Risk, Precision Medicine

## Abstract

**Aims/Hypothesis:** GCK-MODY (Glucokinase-Maturity Onset Diabetes of the Young) causes lifelong, mild hyperglycaemia with high penetrance. Variation in glycaemic phenotype among carriers remains unexplained. We hypothesised that polygenic background contributes to this variability.

**Methods:** To test whether polygenic background contributes to the GCK-MODY clinical phenotype, we analysed polygenic risk scores (PGS) for nine diabetes-related traits in 901 clinically referred individuals with GCK-MODY. We compared these to 7,645 non-diabetic controls and assessed associations between PGS’s and glycaemic measures. Additionally, we evaluated 158 unselected GCK variant carriers from the UK Biobank to examine polygenic effects independent of clinical referral.

**Results:** We observed independent polygenic enrichment for HbA1c (including both glycaemic and non-glycaemic components), fasting glucose, and type 2 diabetes in clinically referred GCK-MODY individuals (0.16-0.33 SD higher, all P < 0.003), but not for type 1 diabetes. In contrast, no such enrichment was seen in *GCK* pathogenic variant carriers from a clinically unselected population-based cohort. In both settings, HbA1c PGSs were associated with measured HbA1c levels in *GCK* carriers (β = 0.91- 0.97, all *P* < 0.009), with effect sizes similar to those in non-carriers. GCK-MODY cases in the top HbA1c quintile had a 3-to-6-fold risk of exceeding the diabetes diagnostic HbA1c threshold (≥ 48 mmol/mol) in clinically selected and clinically unselected cohort respectively.

**Conclusions/interpretation:** Our findings suggest that polygenic background and *GCK* variants interact to modify the glycaemic expression of GCK-MODY, influencing clinical diagnosis despite high penetrance. Our study highlights the importance of integrating both monogenic and polygenic factors to better understand phenotypic variability in monogenic diseases.

**Research in Context:** *What is already known about the subject?:*

- Although GCK-MODY shows high penetrance, individuals vary in their glycaemic phenotype, and the cause of this variability remains unclear.
- Polygenic background has previously been found to modify disease risk and phenotypic variability in other lower penetrant forms of MODY but its contribution to the clinical variability in GCK-MODY is largely unexplored

*What is the key question?:*

- Does polygenic background for diabetes-related traits contribute to variation in the GCK-MODY phenotype?

*What are the new findings?:*

- We identified that clinically referred GCK-MODY cases had an independent enrichment of HbA1c, fasting glucose, and type 2 diabetes polygenic background, potentially increasing the likelihood of clinical referral.
- Higher HbA1c polygenic risk in GCK-MODY was associated with elevated measured HbA1c levels and an increased probability of exceeding the diagnostic threshold for diabetes.

*How might this impact on clinical practice in the foreseeable future?:*

- Polygenic background shapes the clinical expression of GCK-MODY, supporting the integration of monogenic and polygenic information to explain variability in monogenic disease.

## Introduction

There is growing evidence that polygenic background plays a significant role in shaping the clinical expression of monogenic disorders. These disorders arise from rare, large-effect pathogenic mutations, but population studies have shown that penetrance is often lower than expected in unselected cohorts^1–3^. Additionally, clinical presentation can vary widely among family members carrying the same causative variant^4,5^, suggesting the presence of modifying genetic or environmental factors^6^. Polygenic background has been found to influence disease risk and presentation in several monogenic disorders, such as kidney disease, obesity and Long QT syndrome^7–9^. These findings suggest rare pathogenic mutations may interact with an individual’s polygenic background to influence the severity, onset, and clinical trajectory of monogenic diseases.

Polygenic background has been shown to influence disease expression in monogenic diabetes. Maturity Onset Diabetes of the Young (MODY) is the most common form of monogenic diabetes, estimated to contribute to 3.6% of all diabetes cases under 30^10^. Our previous research demonstrated that polygenic background, primarily through type 2 diabetes (T2D) associated pathways, can modify the clinical expression of age-dependent MODY subtypes (*HNF1A*, *HNF1B* and *HNF4A)*^11^. In genetically confirmed cases, polygenic burden was strongly associated with earlier diagnosis, greater severity and explained up to 24% of phenotypic variability^11^. Similar findings in smaller cohorts further highlight the substantial influence of polygenic background on these forms of MODY^4,12^.

However, the extent to which polygenic background shapes clinical expression in highly penetrant GCK-MODY is unclear.

Variability in the GCK-MODY phenotype suggest the potential of polygenic modifiers influencing disease expression. Pathogenic variants in *GCK* are the most common form of monogenic diabetes accounting for up to 60% of MODY cases^13^. GCK-MODY is marked by lifelong fasting hyperglycaemia, with individuals not demonstrating elevated rates of diabetes-related complications and typically not requiring treatment^13–15^. Diagnosis is usually incidental, following routine HbA1c testing in asymptomatic individuals, frequently leading to misdiagnosis as polygenic forms of diabetes and inappropriate glucose-lowering treatment^13^. GCK-MODY has been shown to be highly penetrant, with 96% of carriers having mild hyperglycaemia in the UK Biobank^3^. Despite this, there remains unexplained variability in HbA1c levels in *GCK* carriers (range 38-56 mmol/mol, 5.6-7.3%)^16^, suggesting the presence of polygenic modifiers that may modify disease expression, even in a highly penetrant monogenic disorder.

Understanding how polygenic background contributes to glycaemic variation in GCK-MODY could provide important insights into key biological mechanisms influencing HbA1c levels and glucose regulation. Leveraging the largest clinically referred GCK-MODY cohort to date (n = 901), we used polygenic risk scores for T2D and related metabolic traits to investigate whether polygenic background contributes to and modifies the clinical presentation of GCK-MODY. We replicated our findings in a non-clinically referred cohort of GCK (n = 158) carriers from the UK Biobank.

## Research Design and Methods

### Study Populations

#### Clinically Referred GCK-MODY Cohort

We analysed 901 individuals with genetically confirmed GCK-MODY, referred to the Exeter Genomics Laboratory, Royal Devon University Healthcare NHS Foundation Trust, for monogenic diabetes testing. Referrals were made through routine clinical care based on suspicion of MODY. Clinical characteristics including fasting blood glucose and HbA1c, at time of referral for genetic testing, are detailed in Supplementary Table 1. All biochemistry data was performed locally in NHS laboratories by the referring clinicians. All probands or their guardians provided informed consent, and the North Wales Ethics Committee approved the study.

#### Non-Diabetic Controls

The control group was derived from the Exeter 10,000 study^17^, an ethically approved population-based cohort. Unselected individuals were recruited via general practices across the southwest of the UK. At recruitment, participants completed baseline health questionnaires and provided fasting blood and urine samples for the measurement of metabolic and diabetes-related traits. We included 7,645 individuals who had available genotype data and no evidence of diabetes, defined by having no previous diagnosis of diabetes and a HbA1c ≤ 48 mmol/mol (6.5%)^18^. Participant characteristics are also provided in Supplementary Table 1. Access to samples and genotype data was approved by the NIHR Exeter Clinical Research Facility management committee.

#### UK Biobank Cohort

We used the UK Biobank to assess how polygenic background influences the GCK-MODY phenotype when it is not clinically ascertained. The UK Biobank is a large-scale, prospective population-based study with detailed genetic and phenotype data for approximately 500,000 individuals in the United Kingdom^19^. Participants were aged between 40 to 70 years at recruitment, with recruitment taking place between 2006 and 2010. Detailed phenotypic data was collected through participant questionnaires, interviews and biomarker measurements^19^. We included European 429,491 individuals who underwent both exome sequencing and array genotyping, with their characteristics described in Supplementary Table 2. The UK Biobank resource was approved by the UK Biobank Research Ethics Committee and all participants provided written informed consent to participate.

### Genetic Analysis for *GCK* Variant

#### Exeter MODY Cohort

All GCK-MODY cases included in this study were screened for MODY associated variants using either Sagner sequencing or targeted gene panel testing. These assays were performed by the Exeter Genomics Laboratory, Royal Devon University Healthcare NHS Foundation Trust, as part of routine diagnostic testing, with further methodological details outlined previously^20^. Variant interpretation followed the American College of Medical Genetics and Genomics (ACMG)/Association for Molecular Pathology (AMP) guidelines^21^, and only individuals with *GCK* variants classified as likely pathogenic or pathogenic were included. Variants were annotated using the clinically validated transcript (GenBank: NM_000162.5). Further details on the variant classification protocols used in our local MODY cohort are available in our recent study^3^.

#### UK Biobank

In the UK Biobank, we used exome sequence data to identify carriers of pathogenic *GCK* variants, following the same transcript and classification guideline as above. In this study, we only included protein-truncating variants (PTVs) deemed to be high confidence by the Loss-Of-Function Transcript Effect Estimator (LOFTEE)^22^. To limit the inclusion of false positive low-penetrance variants, we only included missense variants if they were ultra rare in the population (maximum allele count of 2 in gnomAD v2.1.1, MAF <1.4×10-5), been previously seen in a MODY proband (ClinVar or Exeter Genomics Laboratory) and were classified as pathogenic/ likely pathogenic. Sequence read data for all the pathogenic PTV variants was manually reviewed in Integrative Genomics Viewer^23^ to exclude false-positive variants. A full list of pathogenic *GCK* variants identified in the UK Biobank cohort is provided in Supplementary Table 3.

### Array Genotyping

#### GCK-MODY and Non-Diabetic Controls

We used Infinium Global Screening Array genotyping to capture common genetic variation in Exeter MODY cohort and non-diabetes controls. We applied a rigorous quality control pipeline, excluding samples with call rates <98%, sex mismatches, relationship discrepancies, or inbreeding coefficients >0.1. We excluded variants with >2% missingness, minor allele frequency <5%, or if they significantly deviated from Hardy-Weinberg equilibrium (*P* < 1×10^-6^). These procedures were applied both within individual genotyping batches and after batch merging. We performed genotype imputation using the TOPMed reference panel (Version 2)^24^ via the Michigan Imputation Server^25^, using LD-pruned variants as input. Genetic ancestry was determined by principal component analysis (PCA), comparing study participants to reference populations from the 1000 Genomes Project (Phase 3) and the Human Genome Diversity Project^26^, implemented through the GenoPred pipeline (v2.2.1)^27^, with only individuals of European ancestry analysed. Relatedness was assessed using the KING robust algorithm (v2.2.4)^28^. To better model population structure within the cohort, we performed PCA again using FlashPCA (v2.0)^29^. Principal components were first derived from unrelated European individuals and then projected onto related individuals.

#### UK Biobank

UK Biobank individuals were SNP-genotyped using the UK BiLEVE array (∼50,000 individuals), and the UKB Axiom Array (∼450.000 individuals). This dataset underwent extensive central quality control and was imputed to the TOPMed reference panel^24^. In addition, approximately 450,000 individuals underwent exome sequencing using the IDT xGen Exome Research Panel v1.0. The sequencing and quality control pipeline has been described in detail elsewhere^30^. Briefly, variants were called using GATK v3.0, with filters excluding variants with an inbreeding coefficient < –0.03 or lacking at least one genotype meeting the following thresholds: depth (DP) ≥10, genotype quality (GQ) ≥20, and allele balance (AB) ≥0.20 for heterozygotes. For this analysis, we included 429,491 individuals of European ancestry with both array genotyping and exome sequencing data. Ancestry was inferred by principal component analysis using approach applied to our local cohorts.

### Polygenic Score Calculation

To calculate polygenic risk scores (PGS) for nine diabetes related traits, we followed the methodology described in our previous study^11^. In brief, polygenic scores based on genome-wide significant variants were calculated using the plink 1.9’s score function^31^, while genome-wide scores were constructed using the GenoPred pipeline (v 2.2.1)^27^ using the LDpred2 auto model^32^. Weights for fasting glucose and HbA1c genetic risk scores were obtained from Chen *et al.*^33^, using LD-pruned variants (r² < 0.1) from the Trans-Ancestry and European single-ancestry analyses. Overlapping variants from previous T2D association studies (Chen *et al.*^33^, Supplementary Table 4) were classified as T2D increasing if they had been previously found to have a p-value < 0.05 for T2D risk. Similarly, glycaemic and non-glycaemic HbA1c variants were identified using signal classification (Chen *et al*.^33^, Supplementary Table 20). Further details, including the number of variants incorporated and the source studies^33–40^, are provided in Supplementary Table 4.

### Statistical Analysis

To assess whether individuals with GCK-MODY had excess polygenic risk, we tested nine diabetes-related polygenic scores for enrichment compared to controls. All PGSs were standardised to have a mean of zero and a standard deviation of one in the control group. Standardised differences were assessed using linear models adjusted for within-cohort principal components to account for population structure. Due to overlapping genetic variants between some PGSs, each score was initially tested in a separate model. We then performed multivariable logistic regression including all PGSs simultaneously to identify which pathways were independently enriched in GCK-MODY.

To evaluate how polygenic risk influenced the GCK-MODY phenotype, we used mixed-effects models to test the association between polygenic scores and key glycaemic outcomes (fasting glucose and HbA1c). Family ID was included as a random effect to account for within-family correlations among GCK-MODY cases. For control individuals, standard linear models were used as they were unrelated. Each PGS was first assessed in a separate model, adjusting for principal components to account for population structure. PGSs showing significant associations were then included in further models adjusting for relevant clinical and genetic covariates that could influence glycaemic outcomes, including sex, age, BMI, parental history of diabetes and mutation type.

Clinically unselected GCK variant carriers in the UK Biobank enabled us to assess the impact of polygenic background on glycaemic outcomes in a second, population-based setting. We tested associations between each PGS and HbA1c levels, adjusting for available covariates (sex, age, BMI, parental diabetes, mutation type, and genetic principal components). As only the HbA1c PGS showed a significant association, we examined predicted HbA1c levels across PGS percentiles to assess variation in glycaemic levels by polygenic background across carrier status. We also assessed the relationship between HbA1c PGS and the likelihood of exceeding the diagnostic HbA1c threshold for diabetes (≥ 48mmol/mol [6.5%])^18^. These associations were examined using HbA1c PGS by comparing individuals in the top, middle, and bottom PGS quintiles, using both unadjusted and adjusted logistic regression models.

All statistical analyses were performed using R version 4.4.1.

## Results

### Clinically Referred GCK-MODY have Elevated Polygenic Risk for Glycaemic Traits

How factors beyond the primary pathogenic variant influence disease expression in highly penetrant GCK-MODY is not clearly understood. We hypothesis that some of the variation in expression even in this highly penetrance disease could be due to polygenic background. To investigate the contribution of polygenic background to GCK-MODY we analysed polygenic scores for 9 diabetes related traits in 901 clinically referred patients with GCK-MODY (Supplementary Table 1 and 3). As expected, GCK-MODY had higher measured HbA1c and fasting glucose compared to 7645 non-diabetes controls (mean 8.3 mmol/mol and 1.79 mmol/L difference respectively, *P* < 1 ×10^−100^).

Compared to this controls, GCK-MODY patients had significantly higher PGS’s for T2D, HbA1c and Fasting Glucose (0.16 – 0.33 standard deviation increase, all *P*<1×10^-6^) (Figure 1A). Notably, there was no enrichment for T1D PGS (*P* = 0.36). Sensitivity analysis limiting to probands only (N = 709) showed consistent findings (Supplementary Figure 1). The T2D, HbA1c and Fasting Glucose PGS’s were independently enriched after adjusting for other polygenic scores (Figure 1B), indicating that clinical phenotype in clinically referred cases GCK-MODY may be shaped by both GCK variant and glucose-rising biological pathways. To explore this further we stratified the HbA1c and fasting glucose scores by their T2D associations and found that both T2D and non-T2D pathways were enriched in GCK-MODY (Supplementary Figure 2). Similarly, using a partitioned HbA1c risk score, both glycaemic and non-glycaemic pathways were enriched in GCK-MODY (Supplementary Figure 2). These data together suggest that diverse polygenic mechanisms that increased HbA1c contributes to the phonotype of the clinically identified GCK-MODY.

**Figure 1:**
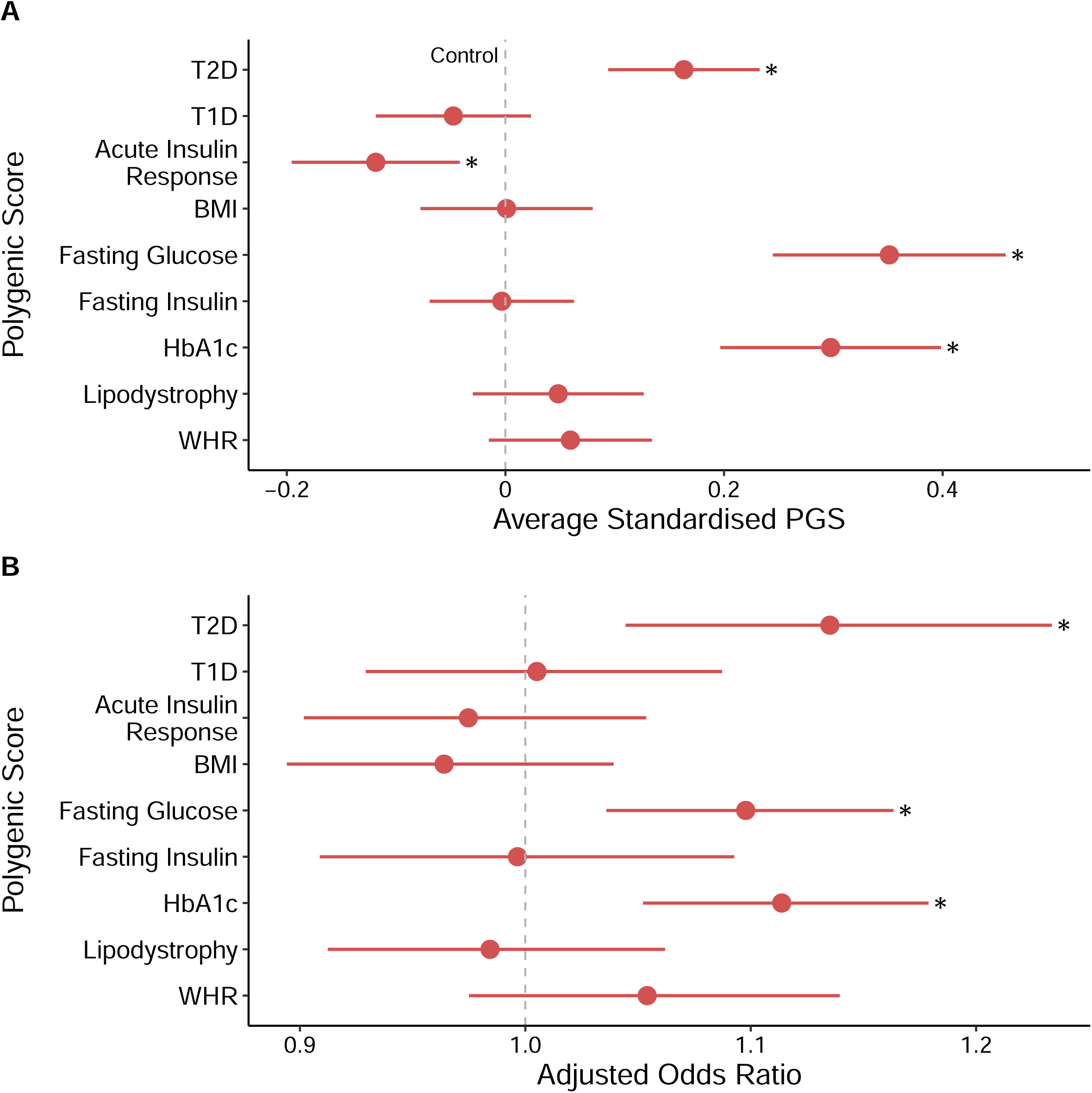
Elevated Polygenic Risk in clinically Referred GCK-MODY. **(A)** Standardised differences in nine diabetes-related polygenic scores, each assessed separately using linear regression, adjusting for the first ten genetic ancestry principal components. GCK-MODY cases (red, N = 901) are compared against control individuals without diabetes (dashed grey line, N = 7,645). **(B)** Adjusted odds ratios for GCK-MODY versus controls, estimated from a multivariable logistic regression model that includes all nine PGS simultaneously, along with the first ten genetic ancestry principal components. All scores are standardised to have a mean of 0 and standard deviation of 1 in controls. Odds ratios represent the change in risk associated with a 1 standard deviation increase in the respective polygenic score. Asterisks denote Bonferroni-adjusted statistically significant differences from controls (*P* < 0.0056). Dots represent the estimates, with error bars indicating 95% confidence intervals. BMI = Body Mass Index, WHR = Waist Hip Ratio.

### Increased Polygenic Risk Modifies HbA1c Levels in Clinically Referred GCK-MODY

Having observed elevated polygenic burden in clinically referred GCK-MODY cases, we next assessed whether this burden influences key features of the GCK-MODY phenotype (HbA1c or Fasting glucose). Among the polygenic scores tested, only the HbA1c PGS showed a significant association with HbA1c levels in GCK-MODY (*P* = 1.8×10^-7^), with a one SD increase in the HbA1c PGS associated with a 0.97 mmol/mol (95% CI: 0.61-1.33 mmol/mol) increase in HbA1c levels (Figure 2A). The effect size was similar to controls (*P* interaction = 0.74). This association remained after adjusting for clinical and genotypic information (β = 0.78, 6.14×10^−7^, Supplementary Table 5). In addition to the effect of HbA1c PGS, we also noted the independent effects of sex (females 1.9 mmol/mol lower HbA1c) and BMI (0.1 mmol/mol increase per kg/m^2^) (Supplementary Table 5).

**Figure 2:**
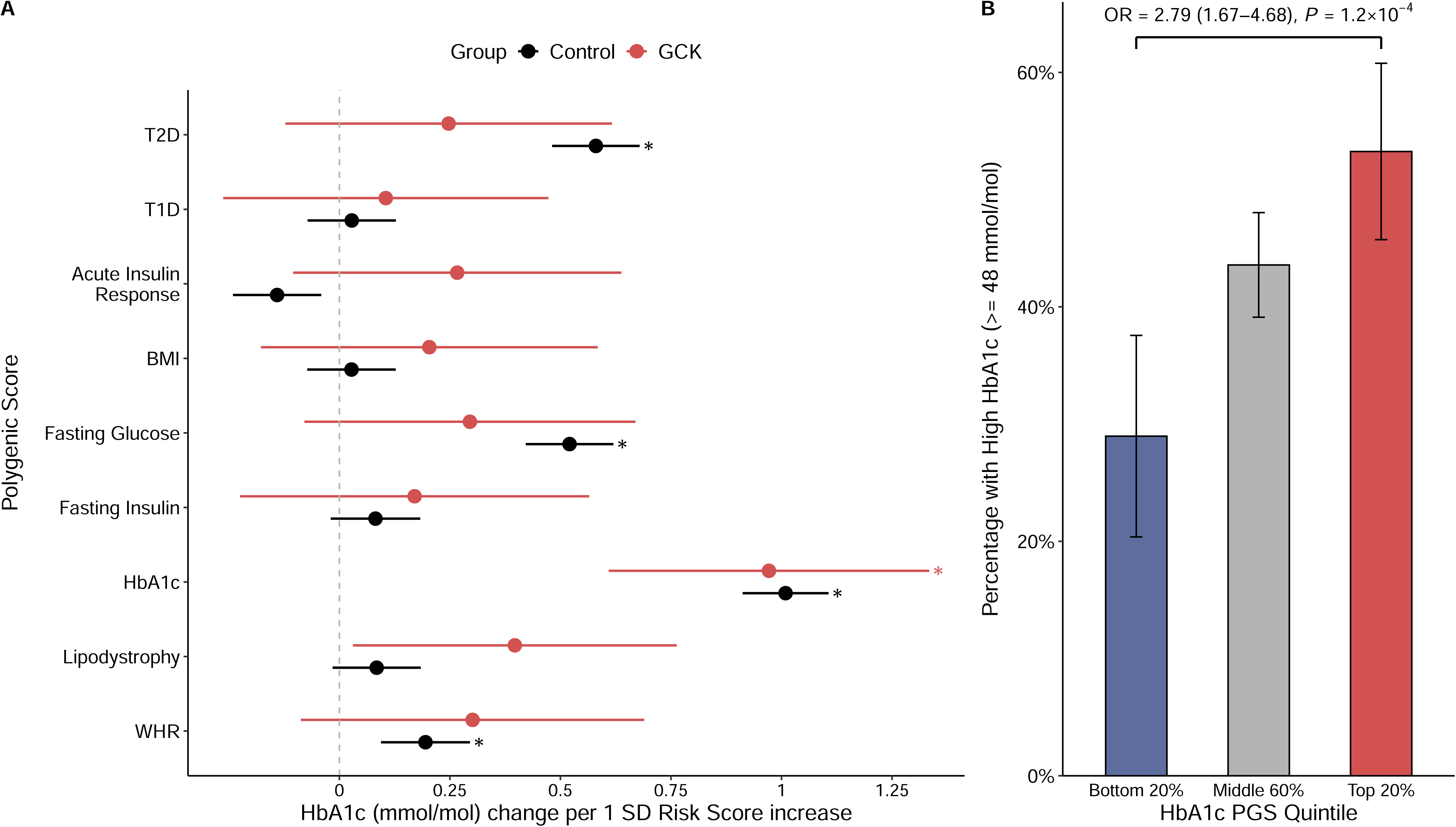
Increased Polygenic Burden Associated with higher HbA1c levels in GCK-MODY. **(A)** Association between polygenic scores for nine diabetes related traits and HbA1c levels (mmol/mol). All scores were assessed individually. For GCK-MODY (red, N= 901), estimates were derived using a mixed-effects linear model with family as a random effect and adjusted for the first ten genetic ancestry principal components. For controls (black, N = 7,645), standard linear regression was used. Estimates represent the effect of a 1 standard deviation increase in the respective polygenic score. Dots represent the estimates, with error bars indicating 95% confidence intervals. Asterisks highlight significant differences after Bonferroni correction (*P* < 0.0028). **(B)** Bar plot showing the percentage of GCK-MODY cases exceeding the diagnostic threshold for diabetes (HbA1c ≥ 6.5%), stratified by HbA1c polygenic score quintiles (bottom 20%, middle 60%, and top 20%). In these groups, 31/109, 207/475, and 90/169 cases, respectively, met the diagnostic threshold. Error bars represent 95% confidence intervals. BMI = Body Mass Index, WHR = Waist Hip Ratio. Odds ratios derived from unadjusted logistic models comparing the top and bottom HbA1c PGS quintile.

Carriers of *GCK* PTVs had significantly higher HbA1c compared to those with missense variants (mean difference = 1.41 mmol/mol, *P* = 2.9×10^-4^) (Supplementary Figure 3, Supplementary Table 5). When partitioned into glycaemic and non-glycaemic components, the HbA1c association in GCK-MODY appeared to be more strongly influenced by the non-glycaemic pathway (β = 0.76, *P* = 4.3×10^−5^), with a similar pattern seen in controls (Supplementary Figure 4). We next examined whether polygenic background was associated with the likelihood of meeting the diagnostic threshold for diabetes (HbA1c ≥ 48 mmol/mol), potentially shaping clinical recognition. In the top HbA1c PGS quintile, 53.3% of GCK-MODY cases exceeded the diagnostic threshold, compared to 29% in the lowest quintile (OR = 2.79, 95% CI: 1.67–4.68, *P* = 1.2×10^−4^) (Figure 2B). These associations remained significant after adjusting for clinical and genetic covariates (Supplementary Table 6). We did not see any significant association between any PGS and fasting blood glucose levels in GCK-MODY (Supplementary Figure 5).

### Polygenic Background Modifies HbA1c Levels in Clinically Unselected *GCK* Carriers

We observed that clinically referred GCK-MODY cases have elevated polygenic burden associated with higher HbA1c levels, which may contribute to clinical recognition. We therefore hypothesized that this enrichment would not appear in unselected *GCK* carriers, although polygenic risk would still influence HbA1c variability. To assess this, we examined individuals carrying pathogenic *GCK* variants (N = 158) from the UK Biobank population-based cohort (N=429,333) (Supplementary Table 2 and 3). Compared to non-carriers, unselected *GCK* carriers showed no enrichment in any of the polygenic scores previously found to be independently elevated in the clinically referred cohort (all *P* > 0.05) (Supplementary Figure 6). However, similar to clinically referred cases, only the HbA1c PGS was significantly associated with HbA1c levels in unselected *GCK* carriers after adjusting for clinical and genetic covariates (Supplementary Figure 7). A one standard deviation increase in HbA1c PGS was associated with a 0.91 mmol/mol increase in HbA1c (0.23 – 1.59 mmol/mol, *P* = 0.009). The effect was similar to non-carriers (*P* interaction = 0.47). Predicted HbA1c values ranged from 43.08 mmol/mol at the 1^st^ percentile (95 CI: 42.02 – 44.03 mmol/mol) to 48.46 mmol/mol at the 99th percentile (95 CI: 47.45 – 49.47 mmol/mol) in *GCK* carriers (Figure 3A). Individuals without *GCK* variants but with the highest polygenic risk did not reach these HbA1c levels (predicted HbA1c = 40.42 mmol/mol, 95% CI: 40.37-40.46 mmol/mol), suggesting that extreme polygenic risk alone cannot fully replicate the glycaemic effect of a pathogenic *GCK* variant as seen in other monogenic disorders. Notably, GCK carriers in the highest HbA1c PGS quintile had significantly greater odds of exceeding the diagnostic threshold for diabetes (HbA1c ≥ 48 mmol/mol) compared to those in the lowest quintile (OR = 5.34, 95% CI: 1.65–17.27, P = 0.005) (Figure 3B). Comparatively, 30.7% of *GCK* carriers (95% CI: 9.1 – 48.5%) in the lowest quintile exceed this threshold, compared to just 4.5% (95% CI: 4.3 - 4.6%) of non-carriers in the highest quintile (*P* = 1.32×10^−5^). These associations remained robust after adjusting for clinical and genetic covariates (Supplementary Table 6), further highlighting how polygenic background and monogenic variants interact to shape disease, even in a highly penetrant disorder.

**Figure 3:**
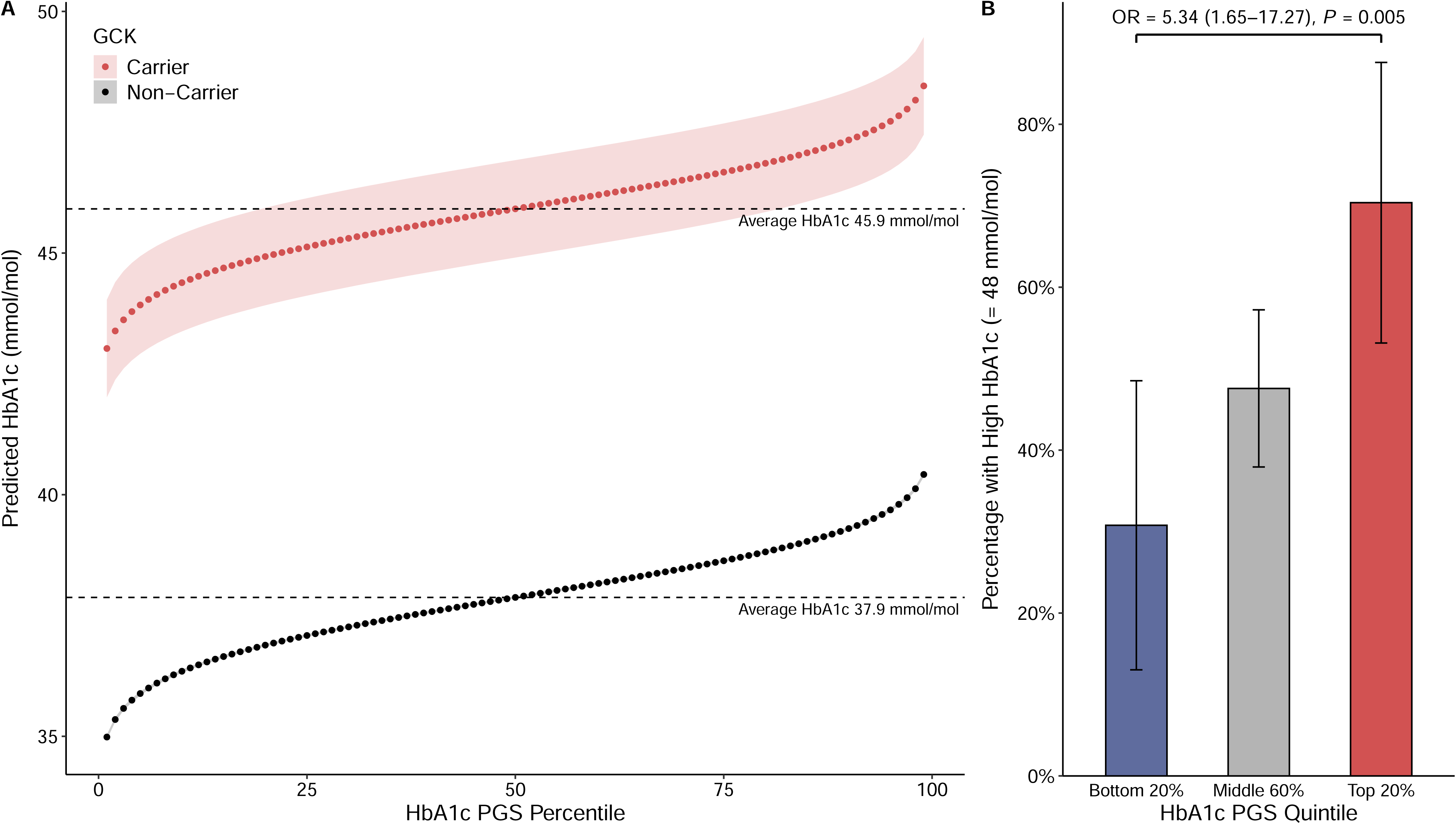
HbA1C PGS modifies phenotype in clinically unselected *GCK* carriers. **(A)** Predicted Hba1c (mmol/mol), assessed using a linear model with HbA1c as a continuous variable adjusted for sex, age, parental history of diabetes, BMI, *GCK* carrier status and mutation type, computing the marginalised effect per percentile. Dots represent percentiles and shaded regions represent 95% confidence intervals. Dashed lines highlight the baseline HbA1c at the 50^th^ percentile for *GCK* carrier (red, N = 158) and non-carriers (black, N = 429,333). **(B)** Bar plot showing the percentage of *GCK* cases exceeding the diagnostic threshold for diabetes (HbA1c ≥ 48 mmol/mol), stratified by HbA1c polygenic score quintiles (bottom 20%, middle 60%, and top 20%). In these groups, 8/26, 49/103 and 19/27 carriers, respectively, exceeded the diagnostic threshold. Error bars represent 95% confidence intervals. Odds ratios derived from unadjusted logistic models comparing the top and bottom HbA1c PGS quintile.

## Discussion

Our analysis reveals that a polygenic background plays a role in shaping the phenotypic expression of highly penetrant GCK-MODY. Using data from 1,059 carriers of pathogenic *GCK* variants, we show that polygenic risk contributes to glycaemic variability, in both clinically referred and population based *GCK* carriers.

Despite GCK-MODY being a highly penetrant monogenic disorder characterised by stable lifelong hyperglycaemia^13^, we observe that polygenic background can influence its clinical expression. Interestingly, our results suggest the polygenic modifiers contributing to GCK-MODY is distinct from other MODY subtypes. In our earlier work^11^, we showed that presentation of progressive, age-dependent forms of MODY (*HNF1A/HNF1B/HNF4A*) are predominately influenced by T2D polygenic risk. In contrast, GCK-MODY patients were independently enriched for T2D, HbA1c and fasting glucose polygenic risk (all *P* < 0.003). Using partitioned HbA1c PGSs, we see that both glycaemic and non-glycaemic pathways contribute to GCK-MODY, highlighting broader biological influences than previously recognised. We observed a lack of enrichment for T1D polygenic risk, consistent with the autoimmune nature of T1D versus the impaired glucose sensing in GCK-MODY^41^, supporting the potential use of T1D risk scores to distinguish GCK-MODY from type 1 diabetes in clinical settings^42^.

This complex interplay between large effect pathogenic mutation and polygenic modifiers supports a liability threshold model of disease. This probability for crossing the threshold for disease expression is primarily driven by the causative pathogenic variant with risk modified by environmental and polygenic factors^43^, now showcased in several monogenic disorders^44^. Since GCK-MODY is present from birth and typically identified later in life from incidental glucose testing^13^, polygenic background may influence whether an individual is identified and referred for genetic testing. This is supported by our finding that clinically referred individuals demonstrated excess polygenic risk, while unselected individuals did not, suggesting polygenic background is a contributing factor to shaping the phenotype that leads to clinical identification and referral. Several studies have demonstrated that increased polygenic risk is associated with increased phenotypic severity, such has in familial epilepsy^45^ and Long-QT Syndrome^7^. While diabetes-related complications are typically considered rare in GCK-MODY patients^15^, polygenic risk could potentially contribute to more severe phenotypes in some cases. For example, a recent report by Ji *et al*^46^ described a *GCK* proband with severe diabetes features, potentially explained by the high insulin resistance polygenic risk enriched on the maternal side. Additionally, *GCK* individuals are often misdiagnosed as polygenic forms of diabetes, leading to unnecessary treatment. This is supported by our findings that *GCK* carriers with higher polygenic risk are more likely to exceed the diagnostic HbA1c threshold (HbA1c ≥ 48 mmol/mol).

This suggests misdiagnosis risk may be amplified by polygenic background, with those with higher polygenic burden more likely to have a phenotype similar to be diagnosed and treated. Further studies are needed to assess the role of polygenic modifiers in the progression and identification of GCK-MODY.

Despite this modifying influence, polygenic background alone was insufficient to replicate the monogenic phenotype of GCK-MODY in the general population. Clinically unselected individuals of high HbA1c polygenic risk (99^th^ percentile, HbA1c: 40.42 mmol/mol) did not display the glycaemic profile seen in *GCK* carriers of low polygenic risk (1^st^ percentile, HbA1c: 43.08 mmol/mol). This differs from conditions like *HNF1A/HNF1B/HNF4A*-MODY^11^ and familial hypercholesterolemia^47^, where high polygenic risk can mimic monogenic phenotypes. The absence of a phenocopy in GCK-MODY could be explained by the particularly high penetrance and strong effect size of the causative mutations. In contrast, recent work by Huerta-Chagoya *et al.*^37^ demonstrated that non-carriers with high polygenic risk had a similar diabetes risk to carriers of the intermediate effect GCK variant p.Val455Glu with low polygenic risk. This suggests large impact pathogenic mutations in GCK-MODY effectively raise the threshold for disease expression, making it difficult for polygenic risk alone to replicate.

This study represents the largest GCK-MODY cohort analysed to date, but several limitations should be acknowledged. Our findings are based on UK-based, European ancestry cohorts, limiting generalisability to other populations. Our clinically selected cohort is derived from routine clinical referrals, however the ascertainment of cases may be influenced by several environmental factors, including healthcare access, socioeconomic status, variability in clinical practices, and other unmeasured confounders. We adjusted our analyses of HbA1c levels for several known or measurable confounding factors, including mutation type, family ID, BMI, age and parental diabetes status. While we have adjusted for known covariates, unmeasured confounders could still potentially affect the associations between PGSs and GCK-MODY phenotype. Notably, we did not observe an association between polygenic background and fasting glucose levels, which may in part reflect variability in the timing and methodology of glucose measurements across different clinical sites, and lower variability explained by the Fasting Glucose PGS^33^. While these limitations remain, the large sample size in the clinically-selected cohort may mitigate some of their effects. Sample size limitations in the unselected GCK-MODY cohort restricted our ability to perform detailed subgroup analyses.

In conclusion, we demonstrate that polygenic background modifies the clinical presentation of highly penetrant GCK-MODY, with HbA1c-related pathways playing a key role in phenotypic variability.

Future work in larger and more diverse cohorts is needed to identify specific modifier variants underlying this polygenic influence. Our findings support integrated approaches combining monogenic and polygenic risk information to enhance biological understanding and improve clinical management of monogenic disease.

## Acknowledgements

The research utilised data from the UK Biobank resource carried out under UK Biobank application number 103356. UK Biobank protocols were approved by the National Research Ethics Service Committee. The work is supported by the National Institute for Health Research (NIHR) Exeter Biomedical Research Centre, Exeter, UK. The Wellcome Trust, MRC and NIHR had no role in the design and conduct of the study; collection, management, analysis, and interpretation of the data; preparation, review, or approval of the manuscript; and decision to submit the manuscript for publication. The views expressed are those of the author(s) and not necessarily those of the Wellcome Trust, Department of Health, NHS or NIHR. For the purpose of open access, the author has applied a CC BY public copyright licence to any Author Accepted Manuscript version arising from this submission.

## Data Availability

The data supporting the findings of this study are available within the article and its supplemental information. The clinical data generated and/or analysed as part of this study are not publicly available because of patient confidentiality and ethical approval associated with the data but are available from the corresponding authors upon reasonable request. The UK Biobank dataset is available from: https://biobank.ctsu.ox.ac.uk

## Author Contributions

Concept, Design or Data Acquisition: K.A.P, M.N.W, A.T.H, K.C

Data curation and analysis: J.M.L, A.M.A, V.K.C, L.N.S, K.A.P All authors contributed to revisions and the final manuscript.

K.A.P is the guarantor of the work, and such had full access to all the data in the study and takes responsibility for the integrity of the data and the accuracy of the analysis.

