## Supplementary Data for "Polygenic Background Contributes to GCK-MODY Clinical Presentation and Glycaemic Variability"

**All Tables and Figures**

| **Characteristics** | **Control** | **GCK-MODY** |
| --- | --- | --- |
| N | 7,645 | 901 |
| Female Sex, n (%) | 4,849 (63.4) | 548 (64.5) |
| Age at Recruitment, y | 54.3 (15.15) | 32.3 (16.4) |
| BMI (kg/m²) | 26.48 (4.68) | 23.5 (4.4) |
| Parent Diabetes, n (%) |  |  |
| None | 6,205 (82.9) | 185 (22.2) |
| Mother | 571 (7.6) | 349 (41.8) |
| Father | 621 (8.3) | 269 (32.2) |
| Both | 84 (1.1) | 32 (3.8) |
| HbA1c (mmol/mol) | 37.8 (4.4) | 46.1 (5.1) |
| Fasting Blood Glucose  (mmol/ L) | 4.99 (0.57) | 6.79 (3.07) |

Supplementary Table 1: Characteristics of GCK-MODY cases and control individuals without diabetes, at referral for genetic testing (GCK-MODY) or recruitment (control). For continuous variables, values are presented as mean (SD), and for categorical variables, counts (n) and percentages (%) are provided. BMI = Body Mass Index, y = years.

| **Characteristics** | **Non-Carrier** | ***GCK*** |
| --- | --- | --- |
| N | 429,333 | 158 |
| Female Sex, n (%) | 233,055 (54.3%) | 84 (53.2%) |
| Age at Recruitment, y | 57.3 (8.0) | 56.6 (8.2) |
| BMI (kg/m²) | 27.4 (4.8) | 27.9 (4.9) |
| Parent Diabetes, n (%) |  |  |
| None | 359,271 (83.7%) | 101 (63.9%) |
| Mother | 33,567 (7.8%) | 31 (19.6%) |
| Father | 32,281 (7.5%) | 20 (12.7%) |
| Both | 4,214 (1%) | 6 (3.8%) |
| HbA1c (mmol/mol) | 38.2 (6.1) | 46.9 (4.9) |
| Fasting Blood Glucose (mmol/L) | 5.1 (1.1) | 6.4 (0.8) |

Supplementary Table 2: Clinical Characteristics of UK Biobank Participants. Clinical characteristics of UK Biobank participants, split by GCK-MODY carriers and noncarriers, at the time of recruitment. For continuous variables, values are presented as mean (SD), and for categorical variables, counts (n) and percentages (%) are provided. BMI = Body Mass Index, y = years.

| **Gene** | **Transcript** | **DNA nomenclature** | **Protein nomenclature** |
| --- | --- | --- | --- |
| *GCK* | NM_000162.5 | c.1358C>T | p.Ser453Leu |
| *GCK* | NM_000162.5 | c.1346C>T | p.Ala449Val |
| *GCK* | NM_000162.5 | c.1322C>T | p.Ser441Leu |
| *GCK* | NM_000162.5 | c.1306A>T | p.Ile436Phe |
| *GCK* | NM_000162.5 | c.1264C>T | p.Arg422Trp |
| *GCK* | NM_000162.5 | c.1228G>C | p.Gly410Arg |
| *GCK* | NM_000162.5 | c.1174C>T | p.Arg392Cys |
| *GCK* | NM_000162.5 | c.1148C>T | p.Ser383Leu |
| *GCK* | NM_000162.5 | c.1142T>C | p.Met381Thr |
| *GCK* | NM_000162.5 | c.1132G>A | p.Ala378Thr |
| *GCK* | NM_000162.5 | c.1019+2T>C | p.? |
| *GCK* | NM_000162.5 | c.1019G>T | p.Ser340Ile |
| *GCK* | NM_000162.5 | c.951C>G | p.His317Gln |
| *GCK* | NM_000162.5 | c.902T>C | p.Leu301Pro |
| *GCK* | NM_000162.5 | c.878T>G | p.Ile293Arg |
| *GCK* | NM_000162.5 | c.878T>C | p.Ile293Thr |
| *GCK* | NM_000162.5 | c.864-1G>A | p.? |
| *GCK* | NM_000162.5 | c.835G>T | p.Glu279* |
| *GCK* | NM_000162.5 | c.823C>G | p.Arg275Gly |
| *GCK* | NM_000162.5 | c.793G>A | p.Glu265Lys |
| *GCK* | NM_000162.5 | c.766G>A | p.Glu256Lys |
| *GCK* | NM_000162.5 | c.748C>T | p.Arg250Cys |
| *GCK* | NM_000162.5 | c.676G>A | p.Val226Met |
| *GCK* | NM_000162.5 | c.667G>A | p.Gly223Ser |
| *GCK* | NM_000162.5 | c.661G>A | p.Glu221Lys |
| *GCK* | NM_000162.5 | c.660C>A | p.Cys220* |
| *GCK* | NM_000162.5 | c.645C>G | p.Tyr215* |
| *GCK* | NM_000162.5 | c.645C>A | p.Tyr215* |
| *GCK* | NM_000162.5 | c.641dup | p.Tyr214* |
| *GCK* | NM_000162.5 | c.626C>T | p.Thr209Met |
| *GCK* | NM_000162.5 | c.601G>T | p.Ala201Ser |
| *GCK* | NM_000162.5 | c.580-13_580-1del | p.? |
| *GCK* | NM_000162.5 | c.580-1G>A | p.? |
| *GCK* | NM_000162.5 | c.579+1G>A | p.? |
| *GCK* | NM_000162.5 | c.571C>T | p.Arg191Trp |
| *GCK* | NM_000162.5 | c.562G>A | p.Ala188Thr |
| *GCK* | NM_000162.5 | c.556C>T | p.Arg186* |
| *GCK* | NM_000162.5 | c.544G>A | p.Val182Met |
| *GCK* | NM_000162.5 | c.540T>G | p.Asn180Lys |
| *GCK* | NM_000162.5 | c.478G>T | p.Asp160Tyr |
| *GCK* | NM_000162.5 | c.478G>A | p.Asp160Asn |
| *GCK* | NM_000162.5 | c.461T>C | p.Val154Ala |
| *GCK* | NM_000162.5 | c.391T>C | p.Ser131Pro |
| *GCK* | NM_000162.5 | c.386G>A | p.Cys129Tyr |
| *GCK* | NM_000162.5 | c.370G>A | p.Asp124Asn |
| *GCK* | NM_000162.5 | c.363+2del | p.? |
| *GCK* | NM_000162.5 | c.351_358del | p.Thr118Aspfs*8 |
| *GCK* | NM_000162.5 | c.316C>T | p.Gln106* |
| *GCK* | NM_000162.5 | c.209-1G>A | p.? |
| *GCK* | NM_000162.5 | c.208+2T>C | p.? |
| *GCK* | NM_000162.5 | c.184G>A | p.Val62Met |

Supplementary Table 3: Pathogenic *GCK* variants identified in the UK Biobank.

| **Trait** | **PubMed ID** | **N SNPs in Score** | **Comments** |
| --- | --- | --- | --- |
| Type 2 Diabetes (T2D) | 38374256 | 1289 | Constructed using plink – score function using genome wide significant variants |
| Type 1 Diabetes (T1D) | 30655379 | 67 | Weighted T1D score using T1DGRS2, available at: https://github.com/t2diabetesgenes/t1dgrs2 |
| Acute Insulin Response | 28490609 | 955764 | Genome-wide polygenic scores, we implemented the GenoPred 2.2.1 pipeline with LDpred2's auto model, which included quality control of summary statistics and genetic data |
| Body Mass Index (BMI) | 25673413 | 886707 |  |
| Fasting Insulin | 34059833 | 1036765 |  |
| Waist Hip Ratio (WHR) | 30239722 | 906879 |  |
| Fasting Glucose | 34059833 | 110  (67 T2D, 34 non-T2D) | Constructed using plink – score function using genome wide significant variants |
| HbA1c | 34059833 | 132  (57 T2D, 54 non T2D)  (35 Glycaemic, 75 non- Glycaemic) |  |
| Lipodystrophy | 27841877 | 53 |  |
| Type 2 Diabetes (T2D)* | 39379762 | 1087858 | Genome-wide polygenic score for T2D, using weights previously derived using PRS- CS, which excluding UKBB participants during testing |

Supplementary Table 4: Polygenic Scores used in analysis. Polygenic scores (PGS) for 9 diabetes related traits used in the analysis. For each trait, the corresponding PubMed ID, and the number of SNPs included in the score are provided. * Used for UK Biobank analysis as it does contain have weights from UK Biobank.

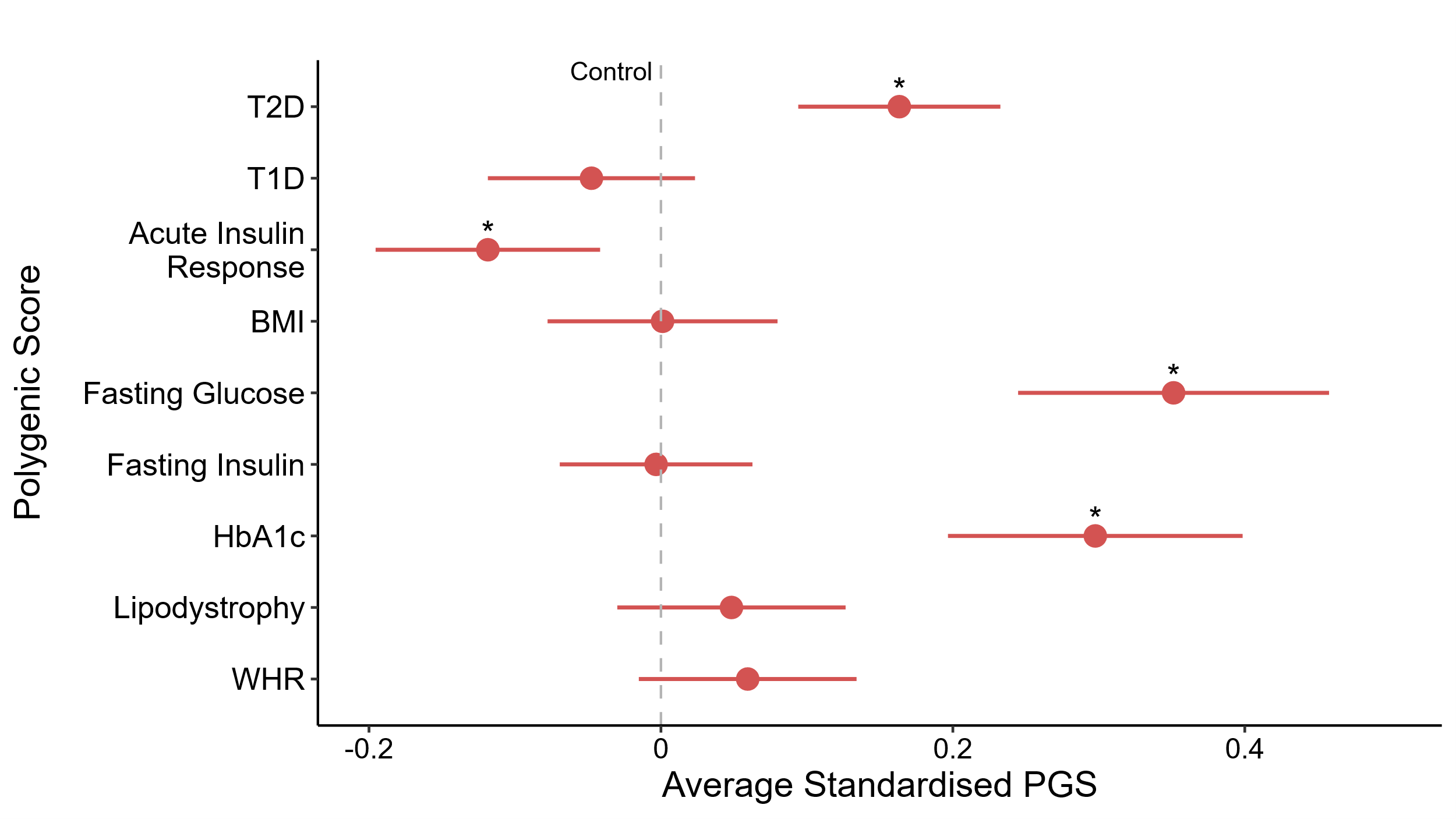

Supplementary Figure 1: Polygenic Risk in clinically referred GCK-MODY probands.

Standardised difference in nine diabetes related polygenic scores, each assed separately using linear regression. GCK-MODY probands (red, N= 709), are compared against control individuals without diabetes (dashed grey line, N = 7,645). All scores are standardised to have a mean of 0 and standard deviation of 1 in controls. Asterisks denote Bonferroni-adjusted statistically significant differences from controls (*P* < 0.0056). Error bars represent 95% confidence intervals. BMI = Body Mass Index, WHR = Waist Hip Ratio.

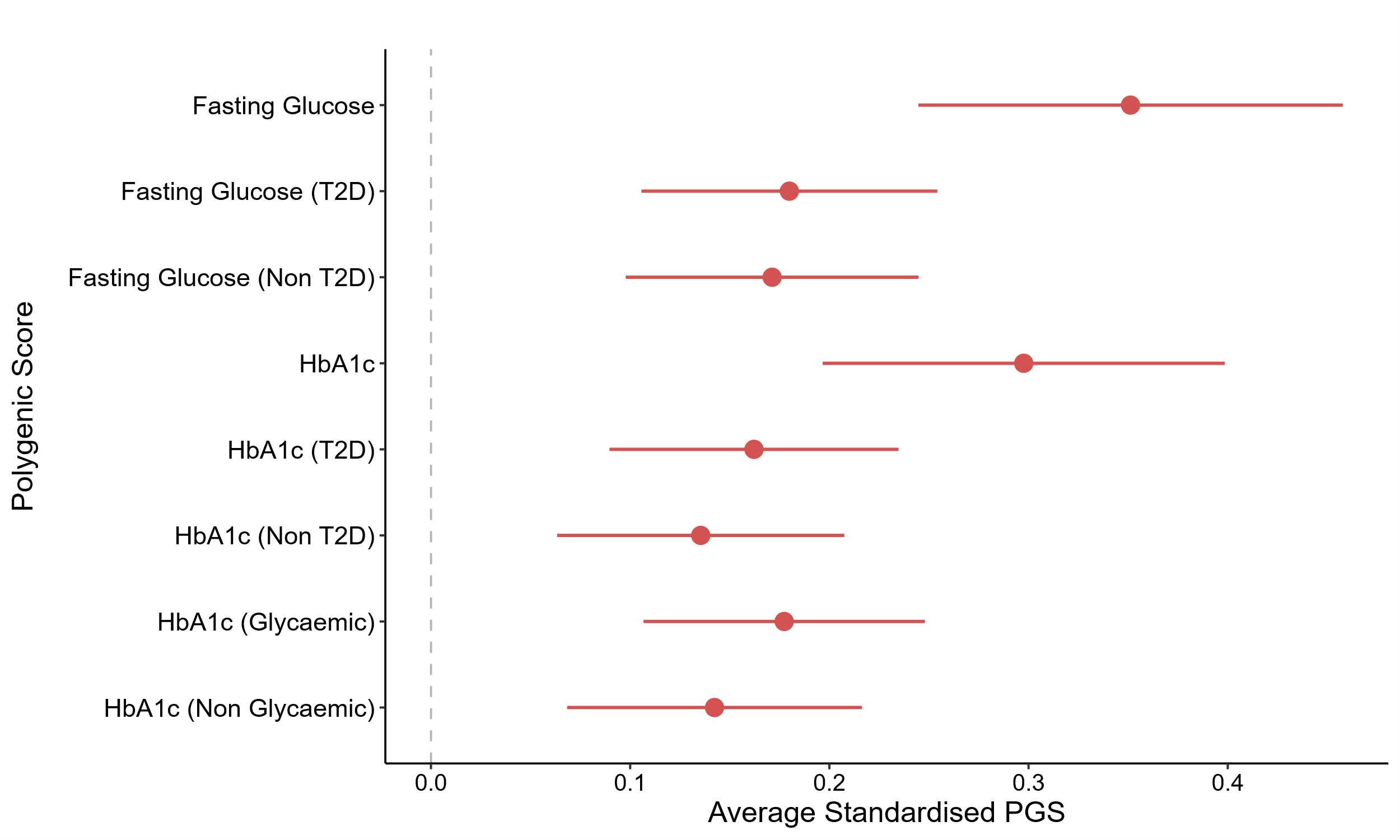

Supplementary Figure 2: Polygenic Risk of Partitioned HbA1c and Fasting Glucose Pathways in GCK-MODY

Standardised difference in Fasting Glucose and HbA1c Polygenic Scores comparing 901 GCK-MODY cases (red) versus 7,645 control individuals without diabetes (dashed grey line). Overlapping genetic variants with previous T2D association studies were identified and used to mark T2D increasing (T2D) or decreasing pathways (Non T2D). Glycaemic and non-glycaemic HbA1c variants were previously identified using signal classification, with more detail in the methods section. All scores are assessed separately using linear regression, adjusting for the first ten genetic ancestry principal components. All scores are standardised to have a mean of 0 and standard deviation of 1 in controls. Error bars represent 95% confidence intervals.

| **Predictor** | **Effect Size**  **(change in HbA1c, mmol/mol)**  **(95% CI)** | ***P*** |
| --- | --- | --- |
| HbA1c PGS (per SD increase) | 0.78 (0.48 – 1.09) | 6.14×10−7 |
| Sex (male vs females) | -1.88 (-2.72 – -1.05) | 1.21×10−5 |
| BMI (per kg/m^2^ increase) | 0.12 (0.03 – 0.21) | 0.008 |
| Mutation Type (Missense vs PTVs) | 1.05 (0.12- 1.98) | 0.02 |
| Parent Diabetes History |  |  |
| Mother | -1.39 (-2.48 – -0.31) | 0.01 |
| Father | -1.18 (-2.22 – -0.15) | 0.03 |
| Both | -1.67 (-3.97 – 0.63) | 0.83 |

Supplementary Table 5: HbA1c PGS increases HbA1c in GCK-MODY cases even after adjusting for clinical and genotype characteristics. Hba1c PGS was included in a mixed effect linear regression model with HbA1c (mmol/mol) as the outcome, and family ID as the random effect. Other covariates included in the model: Sex, Age, BMI, Parental History, Mutation Type and 10 genetic ancestry principal components. All effect sizes in are in DCCT units. Parental Diabetes in reference to subjects whose parents had no history of diabetes. Missense variants, Males and Individuals with no parental history are the reference group in their respective predictor. Protein-truncating variants = PTVs.

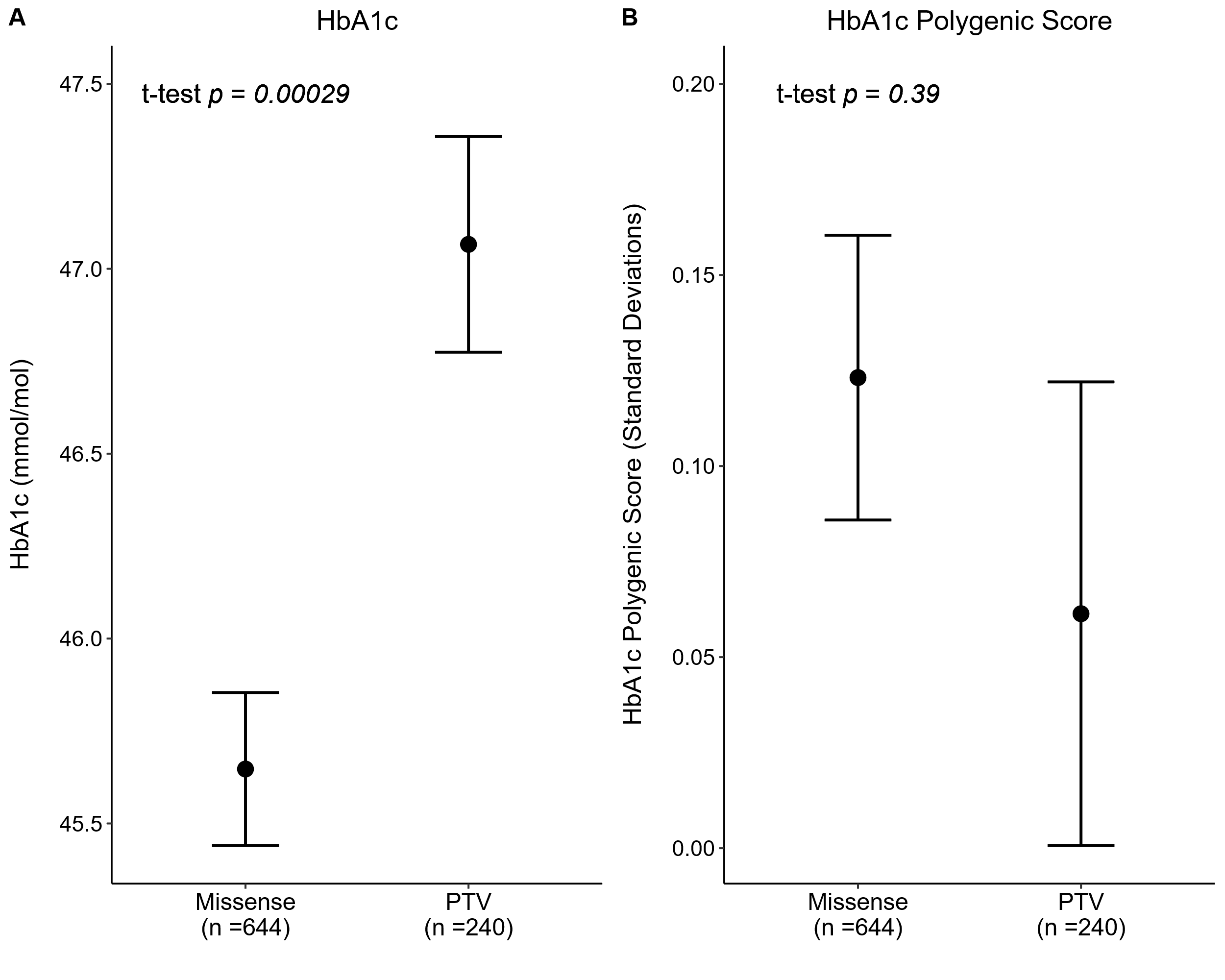

Supplementary Figure 3: HbA1c Effects by Mutations Type in GCK-MODY.

GCK-MODY cases split by predicted effect on protein: Protein Truncting Varaint (PTV) or Missense. (A) Mean Hba1c (mmol/mol) and (B) Mean HbA1c Polygenic Score (Standard Deviations). Points Represent mean values, with error bars represnting 95% confidence intervals. We assesed significance using t-tests. HbA1c polygenic score was standrdised, with controls set to a mean of 0 and standard deviation of 1. Individuals with regulatory and in-frame varaints (N = 17) were excluded in this analysis.

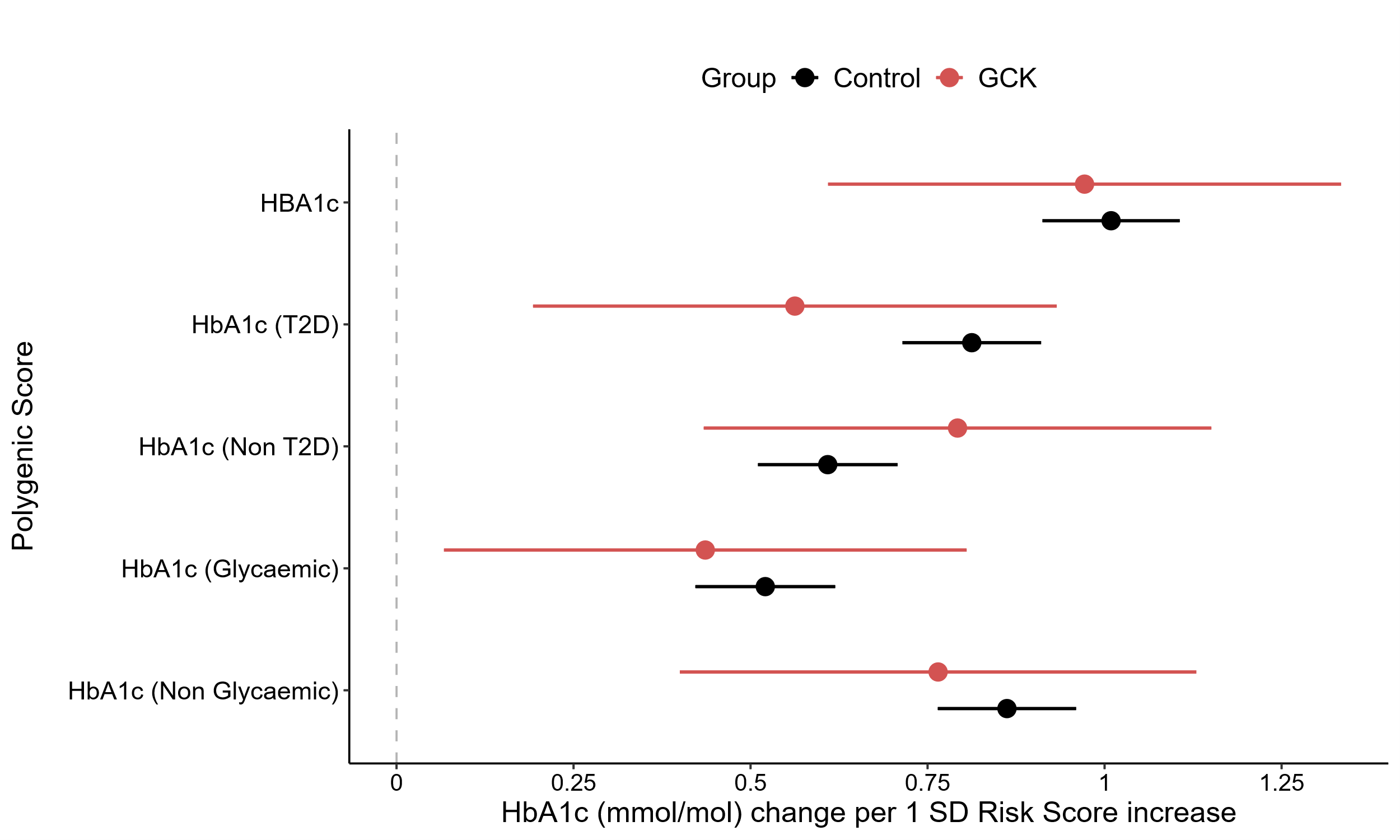

Supplementary Figure 4: Effect of HbA1c partitioned polygenic scores on HbA1c levels

Association between HbA1c partitoned scores and HbA1c levels. Effect sizes reperesent a mmol/mol change in HbA1c per 1 standrad deviation increase in polygenic score. All scores were assesed seperatly. In GCK-MODY (red, N = 901), estimates derived using a mixed-effects linear model with family as a random effect and adjusted for the first ten genetic ancestry principal components. For controls (black, N = 7,645), standard linear regression was used. Dots represent the estimates, with error bars indicating 95% confidence intervals. Overlapping genetic variants with previous T2D association studies were identified and used to mark T2D increasing (T2D) or decreasing pathways (Non T2D). Glycaemic and non-glycaemic HbA1c variants were previously identified using signal classification, with more detail in the methods section. All scores are standardised to have a mean of 0 and standard deviation of 1 in controls.

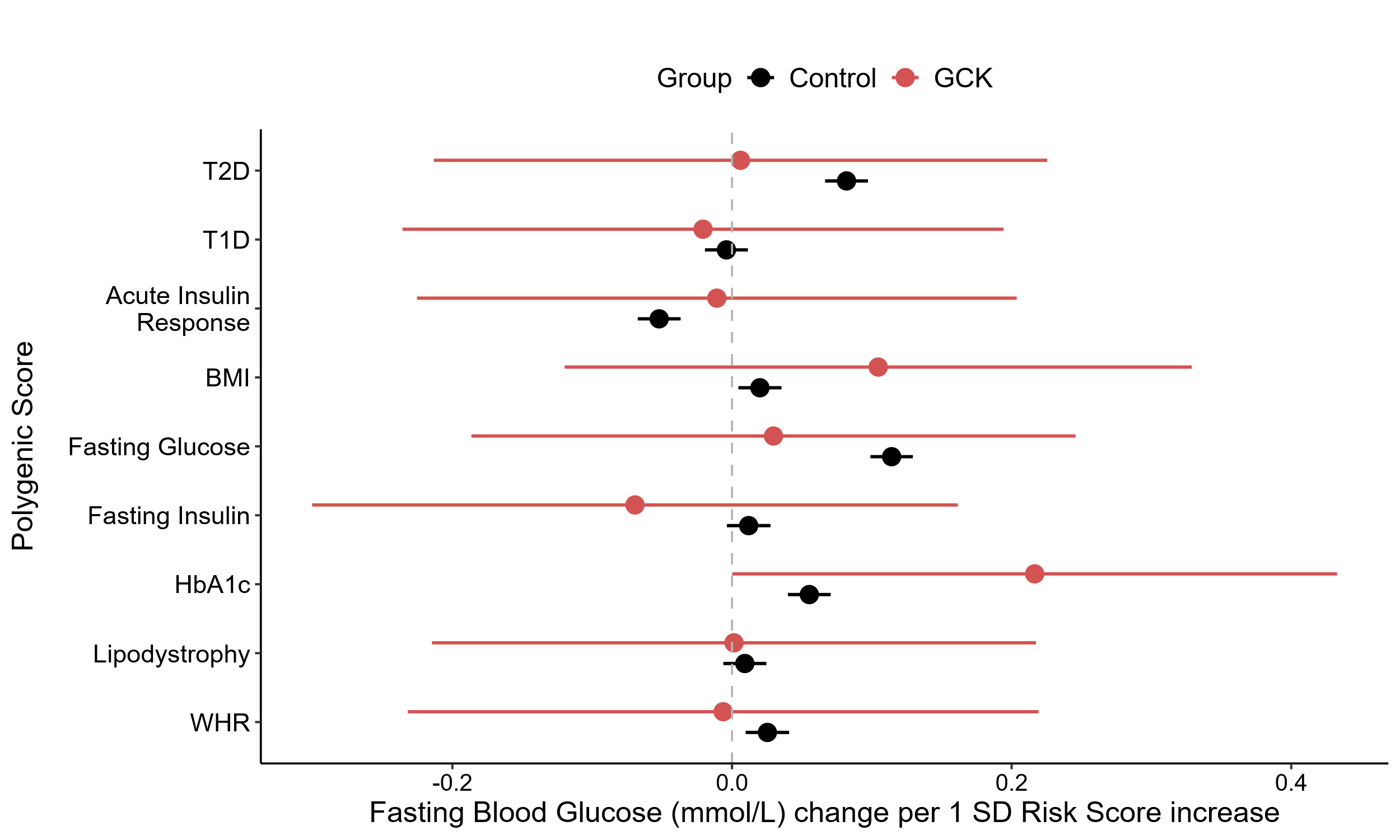

Supplementary Figure 5: Increased Polygenic Burden not Associted with Fasting Glucose levels in GCK-MODY.

Association between polygenic scores for nine diabetes realted traits and Fasting Glucose levels (mmol/L). All scores were assesed individually. For GCK-MODY (red, N= 901), estimates were derived using a mixed-effects linear model with family as a random effect and adjusted for the first ten genetic ancestry principal components. For controls (black, N = 7,645), standard linear regression was used. Estimates represent the effect of a 1 standard deviation increase in the respective polygenic score. Dots represent the estimates, with error bars indicating 95% confidence intervals.

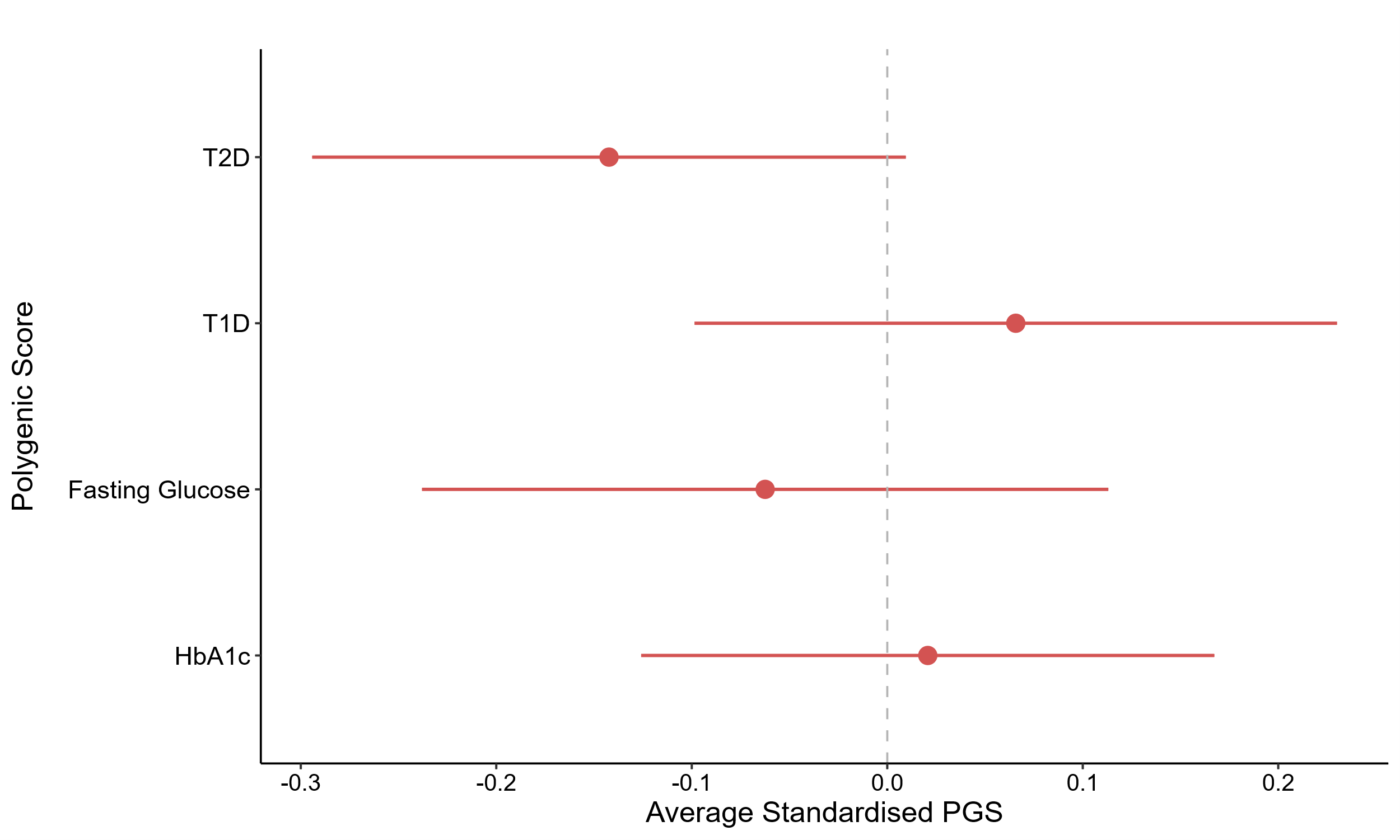

Supplementary Figure 6: No enrichment of polygenic background in clinically unselected *GCK* carriers

Standardized differences in four diabetes-related polygenic scores are shown. These scores were either previously identified as enriched in clinically referred GCK-MODY cases or serve as a negative control (Type 1 diabetes, T1D). Each polygenic score was evaluated separately by comparing 158 *GCK* carriers (red) to 429,333 non-carriers (dashed grey line). Linear regression was used for comparison, adjusting for the first ten principal components within the cohort.

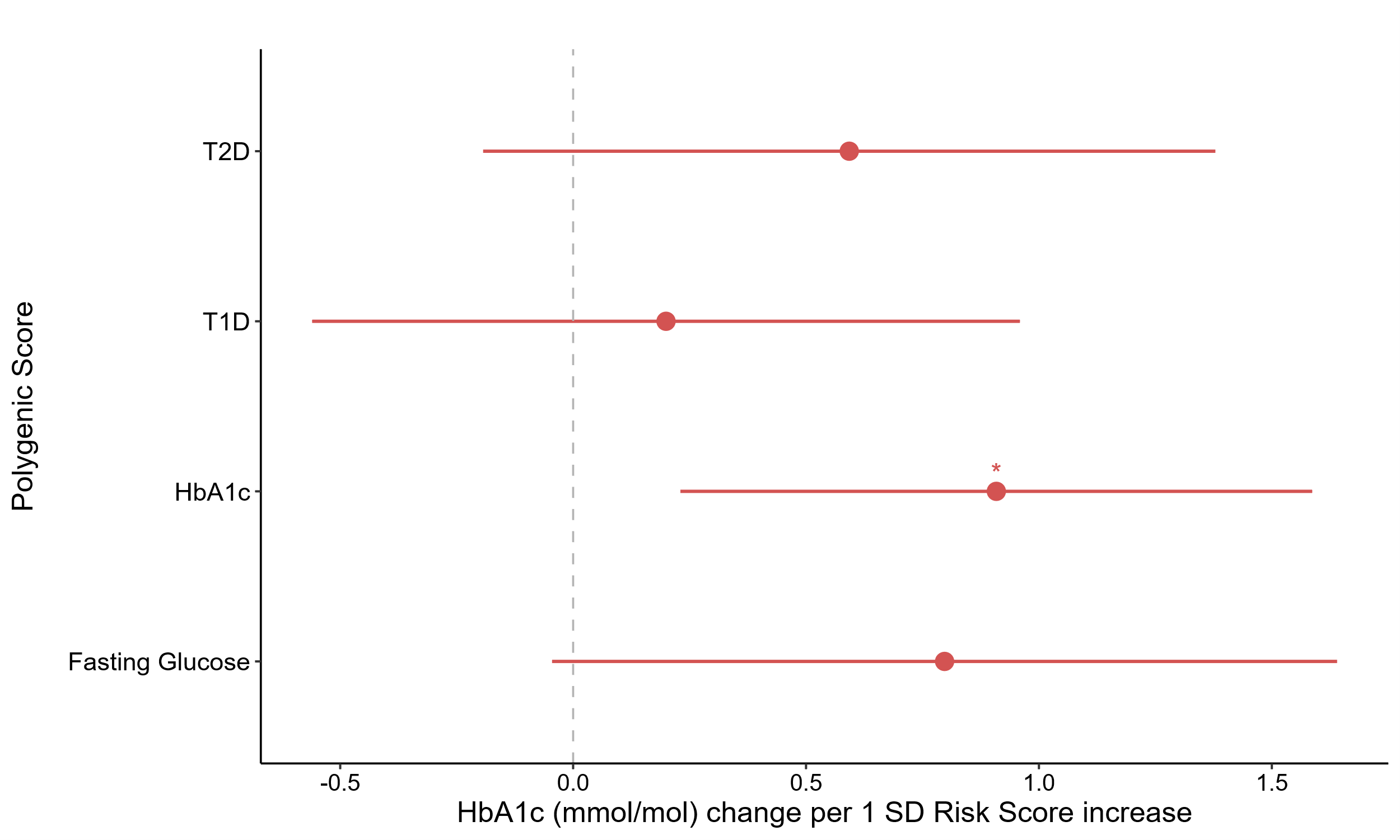

Supplementary Figure 7: Increased Polygenic Burden Associated With higher HbA1c levels in clinically unselected *GCK* Carriers.

Association between polygenic scores for four diabetes related traits and HbA1c levels (mmol/mol). These scores were either previously identified as enriched in clinically referred GCK-MODY cases or serve as a negative control (Type 1 diabetes, T1D). Each score was assessed individually in 158 *GCK* carriers (red), using linear regression models to derive estimates. Estimates represent the effect of a 1 standard deviation increase in the respective polygenic score. Dots represent the estimates, with error bars indicating 95% confidence intervals. Asterisks highlight significant differences after Bonferroni correction (*P* < 0.0125). Covariates included: Sex, Age, BMI, Parental Diabetes, Mutation Type (Protein Truncating Variant or Missense) and the first ten ancestry principal component.

| **Predictor** | **OR (95 CI)**  **Local Cohort** | ***P***  **Local Cohort** | **OR (95 CI)**  **UKBB** | ***P***  **UKBB** |
| --- | --- | --- | --- | --- |
| HbA1c PGS Quintile  Middle 60%  Top 20% | 1.89 (1.2 – 2.99)  2.79 (1.67 - 4.68) | 0.006  1.2×10^−4^ | 2.04 (0.81 – 5.11)  5.34 (1.65 – 17.27) | 0.12  0.005 |
| HbA1c PGS Quintile (Adjusted)  Middle 60%  Top 20% | 2.17 (1.25 -3.76)  3.08 (1.66 -5.76) | 0.006  4.2×10^−4^ | 2.36 (0.84 -6.62)  6.06 (1.59 – 23.03) | 0.1  0.008 |

Supplementary Table 6: Increased HbA1c PGS associated with higher likelihood of exceeding diagnostic diabetes threshold. The likelihood of having an HbA1c level ≥48 mmol/mol in *GCK* carriers, was assessed by stratifying individuals into HbA1c PGS quintiles (bottom 20%, middle 60%, and top 20%). In the local cohort, unadjusted and adjusted mixed-effects logistic models were used, with family-id as a random effect. In the UK Biobank (UKBB), standard logistic regression was applied. Adjusted models included the following covariates, local cohort: sex, age, BMI, parental history of diabetes, mutation type, year of diabetes diagnosis, and 10 genetic ancestry principal components, UKBB: sex, age, BMI, parental history of diabetes, and mutation type, genetic principial components. Odds ratios and 95% confidence intervals are shown for the middle and top quintiles, compared to the bottom quintile.
